# AI-assisted screening of inpatient discharge summaries for potential treatment injury: a single-site service evaluation in a New Zealand district hospital

**DOI:** 10.64898/2026.10.09.26365175

**Authors:** Parag Bhatnagar, William Fearnley

## Abstract

**Objectives:** To assess the feasibility and early performance of an AI-assisted workflow screening inpatient discharge summaries for potential treatment injury requiring clinician review and possible Accident Compensation Corporation (ACC) consideration in New Zealand.

**Design:** Single-site, six-month quality-improvement/service-evaluation pilot using a large language model as an index screening tool with clinician adjudication.

**Setting:** Wairarapa Hospital, New Zealand.

**Participants:** All inpatient discharge summaries produced during the pilot period (n=2,642).

**Interventions:** The model generated structured candidate harm signals with supporting text and certainty labels. Clinicians reviewed all flagged cases and determined whether escalation was appropriate.

**Main outcome measures:** Screening yield, positive predictive value (PPV) for clinician-adjudicated iatrogenic harm, unsupported outputs, overlap with incident reporting and ACC claims, and clinician workload.

**Results:** The workflow generated 237 candidate signals (yield 9.0%, 95% CI 7.9% to 10.1%). Clinicians classified 102 as confirmed iatrogenic harm, 43 as possible harm, 81 as no harm and 11 as unsupported outputs. PPV was 43.0% for confirmed harm (95% CI 36.8% to 49.4%) and 61.2% for confirmed-or-possible harm (95% CI 54.9% to 67.1%); unsupported outputs occurred in 4.6%. Seven ACC claims were lodged following AI-assisted identification. Of 68 organisational ACC claims during the same period, 17 overlapped with AI-identified cases. The workflow required approximately 25–30 minutes of clinician time per week.

**Conclusions:** AI-assisted discharge-summary screening was operationally feasible and increased visibility of potential treatment injuries for clinician review. Further prospective evaluation should use an independent chart-review reference standard to estimate diagnostic accuracy and assess equity, workload and implementation outcomes.

**Registration:** Not applicable.

**Key messages:** *What is already known on this topic:* Healthcare-associated harm is systematically under-detected, with incident reporting and administrative data capturing only a fraction of treatment injuries. Structured record review methods such as the Global Trigger Tool identify substantially higher rates of adverse events but are resource-intensive and difficult to sustain at scale.

*What this study adds:* This study demonstrates that an AI-assisted workflow can feasibly screen inpatient discharge summaries to identify potential treatment injuries for clinician review within routine workflows. The findings suggest that AI can function as a form of visibility infrastructure, surfacing harm signals not consistently captured by existing reporting systems while preserving clinician judgement and governance processes.

*How this study might affect research, practice or policy:* AI-assisted screening could strengthen patient safety surveillance and support more consistent consideration of treatment injury within compensation systems. Future research should evaluate diagnostic accuracy, equity impacts, and workflow design, including whether carefully governed automation of administrative steps can improve escalation processes without displacing clinical oversight.

**Disclaimer:** The AI system generated candidate signals of potential treatment injury only. Final determination of treatment injury status and ACC eligibility remained the responsibility of clinicians and the Accident Compensation Corporation through standard statutory processes.

## 1 Introduction

Healthcare-associated harm remains pervasive yet systematically under-detected across health systems. Despite extensive investment in patient safety initiatives, most hospitals lack reliable mechanisms for identifying adverse events at scale. Voluntary incident reporting systems, while valuable for local learning, are known to capture only a small fraction of clinically significant harm and are subject to reporting bias, professional norms, and time pressure [1,2]. Administrative coding and indicator-based approaches offer additional signals but show heterogeneous validity and limited sensitivity for many harm types [3].

Structured record review methods, such as the Institute for Healthcare Improvement (IHI) Global Trigger Tool [4,5], consistently identify substantially higher rates of adverse events than incident reporting. In comparative studies, trigger-tool review has detected up to ten times more serious adverse events within the same patient populations [1]. However, trigger-tool approaches are labour-intensive, episodic, and difficult to sustain as routine surveillance mechanisms in resource-constrained environments.

New Zealand offers a distinctive context for examining harm detection due to its statutory no-fault injury compensation scheme administered by the Accident Compensation Corporation (ACC). Under the Accident Compensation Act 2001 [6], a treatment injury is personal injury suffered while seeking or receiving treatment from a registered health professional, caused by that treatment, and not a necessary or ordinary consequence given the circumstances. This definition requires clinical interpretation rather than mechanical rule-application.

Unlike tort-based malpractice systems, the ACC scheme separates compensation from blame. While this removes litigation risk, it does not remove the cognitive and administrative burden of recognition and claim initiation. In practice, treatment injury identification is an interpretive act embedded in clinical workflows, professional norms, and organisational priorities.

Discharge summaries represent a pragmatic substrate for harm detection. They are produced for nearly all inpatient episodes and often contain explicit references to complications, adverse drug reactions, procedural difficulties, or unanticipated outcomes. Earlier work using natural language processing demonstrated the feasibility of automated adverse event detection from discharge summaries, but required extensive feature engineering and remained dependent on manual validation [7].

Large language models introduce a qualitatively different capability: they can interpret heterogeneous clinical narratives and generate structured outputs without task-specific rule sets. However, LLMs are probabilistic systems that may produce plausible but unsupported statements, particularly when prompts are weakly constrained. Their application in safety-critical domains therefore requires careful framing, explicit human oversight, and governance mechanisms beyond conventional accuracy metrics.

This study evaluates a pilot implementation of an AI-assisted workflow designed to screen inpatient discharge summaries for potential indicators of treatment injury and escalate these signals to clinicians for review. Rather than automating treatment injury decisions, the intervention functioned as a form of digital visibility infrastructure, surfacing plausible harm signals for clinician review within an existing clinician-governed compensation system.

## 2 Methods

### 2.1 Evaluation framing and DECIDE-AI operationalisation

This work is reported as a quality-improvement/service-evaluation activity and is framed as an early-stage clinical artificial intelligence evaluation, consistent with the objectives and reporting expectations of the DECIDE-AI framework [8]. DECIDE-AI is suited to interventions that are not yet intended for autonomous clinical decision-making, but instead aim to assess safety, feasibility, and integration within real clinical workflows.

The intended use of the AI-assisted workflow was limited to screening and prioritisation. The system was designed to surface candidate treatment-injury signals from discharge summaries for subsequent clinician review. It was not intended to diagnose treatment injury, determine ACC eligibility, or initiate claims. All downstream decisions remained under human control.

Human-AI interaction was deliberately conservative. AI outputs were reviewed asynchronously by clinicians using a structured adjudication template. Reviewers retained full authority to confirm, reject, or defer cases based on clinical judgement and additional context not available to the AI-assisted workflow.

Safety monitoring was incorporated through explicit classification of unsupported or hallucinated outputs. These outputs were treated as safety events rather than model errors and were logged for prompt refinement and governance review.

### 2.2 Setting and data

The pilot was conducted at Wairarapa Hospital (Health New Zealand | Te Whatu Ora). All inpatient discharge summaries for episodes discharged during the preceding week-ending period between July 2025 and January 2026 were included (n = 2,642). No exclusions were applied. The unit of analysis was one discharge summary per inpatient episode. To contextualise the findings, AI-identified treatment injury signals were compared with ACC treatment injury claims lodged during the same period using patient identifiers (NHI) and injury timing to identify overlapping cases. Overlap was assessed manually to determine whether events represented the same injury episode.

### 2.3 AI-assisted workflow architecture and governance

The system used a Microsoft Azure OpenAI Service deployment in Australia Southeast running GPT-4o-mini. All processing occurred within Health New Zealand’s Azure tenant. An Azure App Service acted as a middleware layer, accepting discharge summary text, structuring prompts, invoking the model internally, and returning strictly formatted JSON outputs.

A fixed JSON schema enforced structured outputs and reduced variability. Prompt guardrails required that all identified harms be grounded in explicit discharge summary text, with quoted evidence, and instructed the model not to infer beyond documented information. Unsupported outputs were explicitly classified during human review and treated as safety events.

### 2.4 Human-in-the-loop workflow

Discharge summaries were processed in weekly batches. AI-positive cases were reviewed by clinicians using a structured adjudication template. Each case was classified as confirmed treatment injury, possible treatment injury, no injury, or unsupported/hallucinated output.

Clinician notification followed a structured communication template that explicitly framed AI outputs as advisory and probabilistic. Notifications referenced ACC treatment injury criteria and clarified that escalation decisions remained entirely at the treating clinician’s discretion. All clinicians had been briefed in advance by the Chief Medical Officer regarding the purpose and scope of the pilot.

### 2.5 Rationale for performance metrics

Positive predictive value was selected as the primary performance metric because the AI-assisted workflow operates within a queue-based human review model, where clinician time is the principal limiting resource. In such systems, PPV directly reflects the proportion of reviewed cases that result in clinically meaningful outcomes, and therefore the efficiency of the screening process.

Sensitivity, specificity and false-negative rates were not estimated because non-flagged discharge summaries were not independently reviewed. This omission reflects the early-stage nature of the intervention, where the primary risks relate to false positives, hallucinations and workflow burden rather than comprehensive diagnostic accuracy bench-marking.

### 2.6 Statistical analysis

Descriptive statistics are reported as counts and percentages. Wilson score 95% confidence intervals were calculated for key proportions, including screening yield, positive predictive value, unsupported output rate, escalation rate, overlap with local incident reporting, and overlap with ACC treatment injury claims. Analyses used aggregate counts from the service-evaluation dataset.

### 2.7 Ethics and governance

This study was undertaken and is reported by the authors as a quality-improvement/service-evaluation activity using routinely collected inpatient clinical documentation at Wairarapa Hospital, Health New Zealand | Te Whatu Ora. The AI-assisted workflow generated candidate treatment injury signals for clinician review and did not make autonomous clinical or compensation decisions. Findings are reported in de-identified aggregate form.

Formal ethics committee approval was not obtained before the evaluation commenced. Subsequent advice was sought from Health New Zealand research governance; however, the correspondence available to the authors does not constitute a formal ethics committee approval or waiver.

### 2.8 Patient and public involvement

Patients and members of the public were not involved in the design, conduct, reporting or dissemination planning of this quality-improvement/service-evaluation activity. The activity used routinely collected operational clinical documentation and focused on workflow feasibility and governance. Future prospective evaluations should include patient and whānau perspectives, particularly in relation to transparency, equity, and access to ACC treatment injury entitlements.

## 3 Results

Over the six-month pilot period, 2,642 inpatient discharge summaries were screened. No exclusions were applied. The AI-assisted workflow flagged 237 discharge summaries as containing potential treatment injury signals, corresponding to a screening yield of 9.0% (237/2,642; 95% CI 7.9% to 10.1%). Flagged cases were forwarded to clinician review in weekly batches.

All AI-flagged cases underwent structured clinician adjudication. Of the 237 flagged cases, 102 (43.0%) were classified as confirmed treatment injury, 43 (18.1%) as possible treatment injury where available documentation was insufficient for confirmation, 81 (34.2%) as no treatment injury, and 11 (4.6%) as unsupported or hallucinated outputs. The positive predictive value for confirmed treatment injury was 43.0% (95% CI 36.8% to 49.4%). When confirmed and possible injuries were combined, PPV increased to 61.2% (95% CI 54.9% to 67.1%).

**Table 1.** Clinician adjudication outcomes for AI-flagged cases. Confidence intervals are Wilson score 95% confidence intervals for proportions.

| Outcome | Count | % of AI-flagged cases | 95% CI |
| --- | --- | --- | --- |
| Confirmed treatment injury | 102 | 43.0 | 36.8 to 49.4 |
| Possible treatment injury | 43 | 18.1 | 13.8 to 23.5 |
| No treatment injury | 81 | 34.2 | 28.4 to 40.4 |
| Unsupported/hallucinated output | 11 | 4.6 | 2.6 to 8.0 |
| Total | 237 | 100.0 | - |

The AI-assisted workflow assigned a triage label reflecting estimated likelihood of treatment injury. Performance differed substantially by label. Among cases labelled very likely, 78.2% were confirmed as treatment injury following clinician review. In contrast, 25.5% of cases labelled possible were confirmed. All unsupported or hallucinated outputs occurred within the possible category. This stratification enabled pragmatic thresholding to align review effort with available clinical capacity.

Of the 102 clinician-confirmed treatment injuries, 58 (56.9%; 95% CI 47.2% to 66.1%) were escalated for ACC consideration. The remaining 44 (43.1%) were not escalated. Reasons for non-escalation included clinician determination that the outcome represented an ordinary or expected consequence of treatment, identification of existing ACC claims already lodged, insufficient documentation to support claim initiation, judgement that escalation would not provide meaningful patient benefit, manual workflow errors where escalation was not sent to the senior clinician responsible for lodging treatment injury claims, and cases where agreement to lodge a claim was not followed by formal submission within the ACC system.

Across the pilot, seven new ACC treatment injury claims were lodged directly as a result of AI-assisted identification. At the time of analysis, three claims had been accepted, two had been declined, and two were pending or on hold. At the time of reporting, accepted claims had reimbursements totalling NZD 3,373.57, while additional amounts remained pending determination.

**Table 2.** Selected workflow, safety and ACC comparison metrics. These descriptive figures are not sensitivity or incidence estimates because non-flagged summaries were not independently reviewed.

| Metric | Count; percentage (95% CI) | Interpretation |
| --- | --- | --- |
| Screening yield | 237/2,642; 9.0% (7.9 to 10.1) | Proportion of discharge summaries flagged for review |
| Confirmed treatment injury PPV | 102/237; 43.0% (36.8 to 49.4) | Confirmed harm among AI-flagged cases |
| Confirmed-or-possible PPV | 145/237; 61.2% (54.9 to 67.1) | Confirmed or possible harm among AI-flagged cases |
| Unsupported output rate | 11/237; 4.6% (2.6 to 8.0) | Flagged cases with unsupported/hallucinated rationale |
| Escalation for ACC consideration | 58/102; 56.9% (47.2 to 66.1) | Confirmed injuries escalated for ACC consideration |
| Incident-reporting overlap | 21/102; 20.6% (13.9 to 29.4) | Confirmed injuries captured in local incident reporting |
| AI signals corresponding with lodged ACC claims | 17/145; 11.7% (7.5 to 18.0) | Confirmed-or-possible AI signals overlapping with ACC claims |
| ACC claims detected by AI workflow | 17/68; 25.0% (16.2 to 36.4) | ACC treatment injury claims also detected by AI screening |

Among the 102 confirmed treatment injuries, only 21 (20.6%; 95% CI 13.9% to 29.4%) were identified as having been captured through the local incident reporting system. The remaining 81 (79.4%; 95% CI 70.6% to 86.1%) had no corresponding incident report recorded.

Overlap with administrative coding was heterogeneous. Coding clearly reflected treatment injury in a minority of cases, was partially indicative in some, and absent or indeterminate in others. This variability limited the utility of coding data as a standalone comparator but suggested that AI-assisted screening identified cases not reliably visible through existing mechanisms.

During the pilot period (13 July 2025 to 13 January 2026), the AI-assisted workflow identified 145 confirmed or possible treatment injury signals from inpatient discharge summaries. ACC records show 68 treatment injury claims lodged with an injury date within the same timeframe. Reconciliation of the two datasets identified 17 injury events appearing in both datasets. These cases included both injuries escalated through the AI-assisted workflow and injuries lodged independently through routine clinical processes. Overall, 128 potential treatment injuries were identified by the AI workflow but were not lodged as ACC claims, and 51 ACC treatment injury claims were not detected by the AI screening tool. These figures indicate that 11.7% of AI-identified confirmed-or-possible signals corresponded with lodged ACC claims, while 25.0% of ACC claims were detected by the AI screening workflow. The relationship between the two datasets is shown in Figure 1.

**Figure 1.**
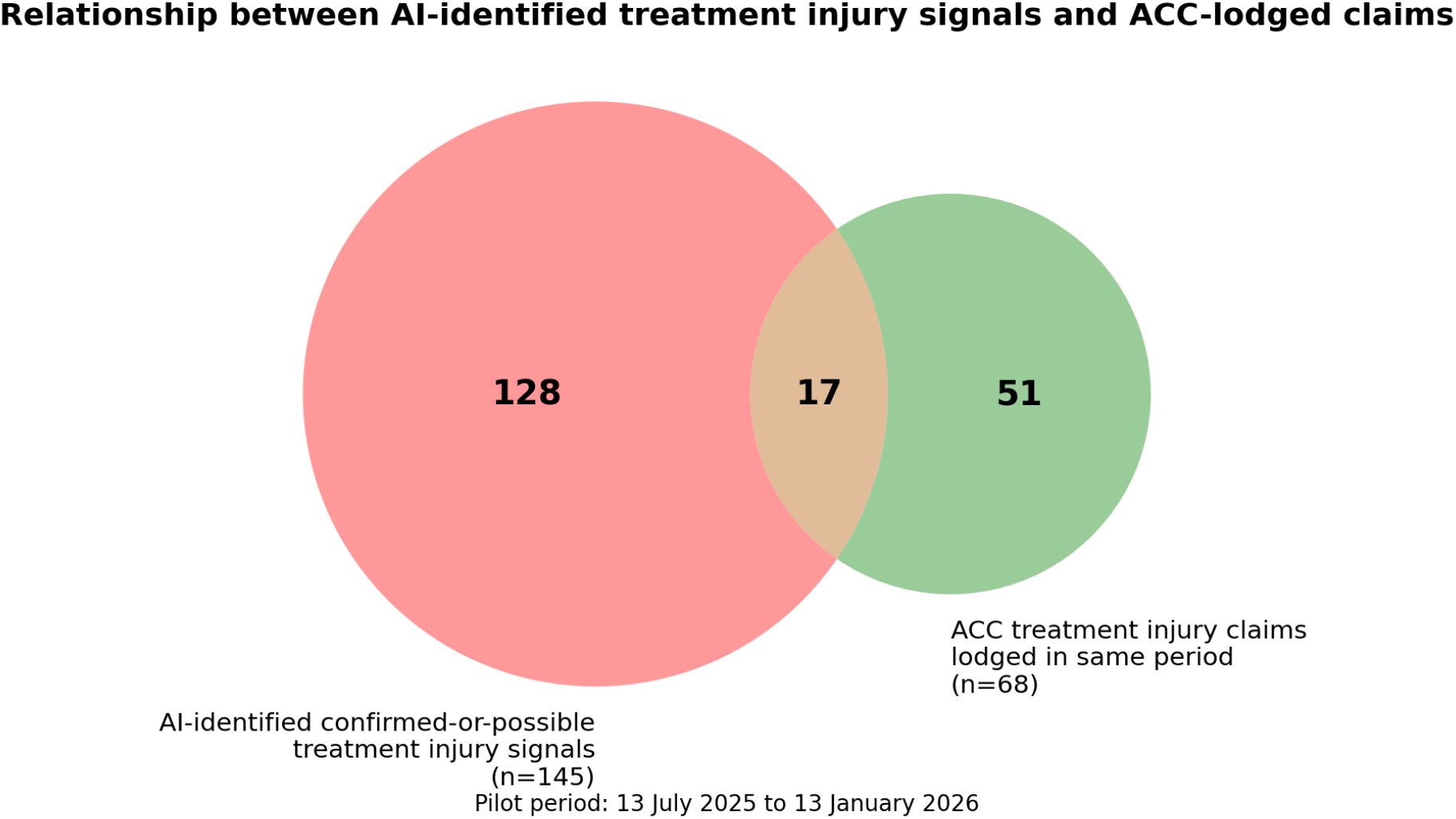
Relationship between AI-identified confirmed-or-possible treatment injury signals and ACC-lodged treatment injury claims during the pilot period, 13 July 2025 to 13 January 2026. The overlap represents 17 documented injury events appearing in both datasets after reconciliation using National Health Index identifiers and injury timing.

Operational workload associated with the AI-assisted workflow was modest. Weekly discharge summary report generation required approximately two minutes. Each weekly batch produced an average of ten AI-positive cases. Manual clinical review of flagged cases required approximately 20 minutes per week, with an additional 5 to 10 minutes devoted to ACC-related follow-up activities. Overall, the workflow required approximately 25 to 30 minutes of clinician time per week.

## 4 Discussion

This pilot demonstrates that LLM-assisted screening of discharge summaries can feasibly surface plausible treatment injuries for structured clinician review within a no-fault compensation context. The central contribution is not algorithmic performance alone, but the integration of AI into a socio-technical system of safety, compensation, and governance.

The partial overlap between AI-identified cases and ACC-lodged claims reflects the different mechanisms through which treatment injury events become visible within healthcare systems. The AI workflow operates prospectively on discharge documentation to surface potential indicators of iatrogenic harm, whereas ACC claims depend on clinician judgement, eligibility criteria and statutory compensation processes. The two datasets therefore represent overlapping but distinct pathways of harm recognition rather than interchangeable measures of injury incidence.

The governance model extended beyond technical safeguards to include structured clin-ician communication. AI-flagged cases were accompanied by non-directive notifications emphasising clinician discretion and alignment with ACC eligibility criteria. This framing was critical to maintaining trust and preventing misinterpretation of the system as an automated claims trigger.

Most confirmed treatment injuries identified in this pilot were not captured through incident reporting, consistent with international evidence that voluntary reporting under-ascertains harm. The AI-assisted workflow functioned as a visibility amplifier, drawing clinician attention to cases that might otherwise remain unexamined. Importantly, detection did not equate to escalation. Many confirmed injuries were not pursued as ACC claims due to eligibility judgement or contextual factors. This attrition is not a failure of the system, but a reflection of clinical and administrative realities.

Improving consistency in treatment injury consideration may have equity implications. Under-lodgement may disproportionately affect patients who are less able to advocate for themselves. While this study did not assess demographic effects, the workflow provides a foundation for more equitable harm recognition. This claim is exploratory and requires empirical testing.

Although ACC is unique, the findings have broader relevance. In tort-based systems, similar AI-assisted screening could support internal safety review, risk management, or disclosure processes. The key insight is that AI’s value lies in supporting human judgement, not replacing it.

Hallucinations, while infrequent, underscore the necessity of strong governance. Unsupported outputs should be treated as safety signals requiring monitoring, prompt refinement and audit. Scaling such systems without explicit governance risks eroding trust. Compared with structured trigger-tool review, the AI-assisted approach described here requires substantially less clinician time, operating continuously rather than episodically. Relative to earlier NLP-based adverse event detection systems, the use of LLMs enabled broader linguistic coverage without task-specific feature engineering, but introduced new safety risks that require explicit governance.

### 4.1 Limitations and future research

This evaluation did not assess sensitivity or missed cases. In addition, attribution of ACC claim lodgement to the AI-assisted workflow cannot be determined with certainty for all overlapping cases because treatment injury claims may also arise through routine clinical processes. Discharge summaries provide incomplete context, inflating the possible category. Results are single-site. Future work should adopt a hybrid effectiveness-implementation design [9] incorporating reviewed negative samples, richer documentation, patient and whānau perspectives, and longitudinal outcome assessment.

### 4.2 Conclusion

AI-assisted screening of discharge summaries can enhance the visibility of potential treatment injuries within a no-fault compensation system, supporting clinician review, equity and learning. Its value lies in infrastructure and governance, not automation. Careful evaluation and oversight are essential for safe scale-up.

The marked discrepancy between AI-identified positive signals and the relatively small number of claims ultimately materialised highlights the complexity of treatment injury recognition within a clinician-governed compensation framework. This gap does not imply over-detection by the AI-assisted workflow; rather, it reflects procedural, cognitive and administrative factors that shape escalation decisions. Future work should investigate whether carefully designed workflow supports, including limited and well-governed automation of administrative steps, could improve the consistency and equity of treatment injury consideration while preserving clinician judgement and statutory safeguards.

## Data Availability

All data produced in the present study are available upon reasonable request to the authors.

## Statements and declarations

### Patient consent for publication

The manuscript reports de-identified aggregate findings and contains no identifiable patient-level information. Individual patient consent for publication was not sought. This statement does not constitute a waiver of research ethics review.

### Data availability statement

Patient-level clinical data are not available for sharing because they contain sensitive health information. De-identified aggregate data supporting the reported results may be available from the corresponding author on reasonable request, subject to Health New Zealand governance, privacy and data-access approvals.

### Funding

No specific funding was received for this study.

### Competing interests

The authors declare no competing interests.

### Author contributions

PB and WF contributed to study conception, interpretation of findings, and manuscript preparation. PB led AI/automation workflow analysis, implementation framing and manuscript drafting. WF led clinical oversight, treatment-injury adjudication and interpretation of ACC treatment injury processes. Both authors reviewed and approved the manuscript. PB is the guarantor.

### Transparency statement

The guarantor affirms that the manuscript is an honest, accurate and transparent account of the activity being reported; that no important aspects of the activity have been omitted; and that any discrepancies from the activity as planned have been explained.

### Use of artificial intelligence

OpenAI ChatGPT was used during manuscript preparation to assist with language editing, restructuring, reference-format conversion and document formatting. The authors reviewed and revised all AI-assisted output, verified the underlying study data and references, and take full responsibility for the accuracy and integrity of the submitted manuscript. This editorial use is distinct from the GPT-4o-mini-based clinical workflow evaluated in the study, which is described in the Methods.

### Reporting guideline

Reporting was informed primarily by DECIDE-AI for early-stage clinical AI evaluation [8], with SQUIRE 2.0 used where relevant to the quality-improvement/service-evaluation context [10]. A completed DECIDE-AI reporting crosswalk is supplied as a separate submission file.

### Dissemination plans

Findings are intended for dissemination through peer-reviewed publication and internal Health New Zealand clinical governance and digital health forums. Future prospective evaluations should include patient and whānau-facing dissemination planning.

## Notes

### Competing Interest Statement

The authors are practitioners and report this work as part of the quality improvement exercise undertaken.

### Author Declarations

This study was undertaken and is reported by the authors as a quality-improvement/service-evaluation activity using routinely collected inpatient clinical documentation at Wairarapa Hospital, Health New Zealand | Te Whatu Ora. The AI-assisted workflow generated candidate treatment injury signals for clinician review and did not make autonomous clinical or compensation decisions. Findings are reported in de-identified aggregate form. Formal ethics committee approval was not obtained before the evaluation commenced. Subsequent advice was sought from Health New Zealand research governance; however, the correspondence available to the authors does not constitute a formal ethics committee approval or waiver. The patient datasets, including the inpatient discharge summaries, were de-identified prior to their use in this study.

## References

1. Classen DC, Resar R, Griffin F, Federico F, Frankel T, Kimmel N, et al. ‘Global trigger tool’ shows that adverse events in hospitals may be ten times greater than previously measured. Health Aff (Millwood). 2011;30(4):581–589. doi:10.1377/hlthaff.2011.0190.

2. Hibbert PD, Molloy CJ, Schultz TJ, Carson-Stevens A, Braithwaite J. Comparing rates of adverse events detected in incident reporting and the Global Trigger Tool: a systematic review. Int J Qual Health Care. 2023;35(3):mzad056. doi:10.1093/intqhc/mzad056. PMID:37440353.

3. Maass C, Kuske S, Lessing C, Schrappe M. Are administrative data valid when measuring patient safety in hospitals? A comparison of data collection methods using a chart review and administrative data. Int J Qual Health Care. 2015;27(4):305–313. doi:10.1093/intqhc/mzv045 PMID:26133382.

4. Classen DC, Lloyd RC, Provost L, Griffin FA, Resar R. Development and evaluation of the Institute for Healthcare Improvement Global Trigger Tool. J Patient Saf. 2008;4(3):169–177. doi:10.1097/PTS.0b013e318183a475.

5. Griffin FA, Resar RK. IHI Global Trigger Tool for Measuring Adverse Events. 2nd ed. Cambridge, MA: Institute for Healthcare Improvement; 2009.

6. Accident Compensation Act 2001 (NZ), s 32 (treatment injury). New Zealand Legislation.

7. Hripcsak G, Bakken S, Stetson PD, Patel VL. Automated detection of adverse events using natural language processing of discharge summaries. J Am Med Inform Assoc. 2005;12(4):448–457. doi:10.1197/jamia.M1794.

8. Vasey B, Nagendran M, Campbell B, et al. Reporting guideline for the early-stage clinical evaluation of decision support systems driven by artificial intelligence: DECIDE-AI. Nat Med. 2022;28(5):924–933. doi:10.1038/s41591-022-01772-9. PMID:35585198.

9. Curran GM, Bauer M, Mittman B, Pyne JM, Stetler C. Effectiveness-implementation hybrid designs: combining elements of clinical effectiveness and implementation research to enhance public health impact. Med Care. 2012;50(3):217–226. doi:10.1097/MLR.0b013e3182408812 PMID:22310560.

10. Ogrinc G, Davies L, Goodman D, Batalden P, Davidoff F, Stevens D. SQUIRE 2.0: revised publication guidelines from a detailed consensus process. BMJ Qual Saf. 2016;25(12):986–992. doi:10.1136/bmjqs-2015-004411. PMID:26369893.

